# Identification of metabolomics biomarkers for type 2 diabetes: triangulating evidence from longitudinal and Mendelian randomization analyses

**DOI:** 10.1101/2020.10.30.20222836

**Authors:** Eleonora Porcu, Federica Gilardi, Liza Darrous, Loic Yengo, Nasim Bararpour, Marie Gasser, Pedro Marques Vidal, Philippe Froguel, Gerard Waeber, Aurelien Thomas, Zoltán Kutalik

## Abstract

The number of people affected by Type 2 Diabetes Mellitus (T2DM) is close to half a billion and is on a sharp rise, representing a major and growing public health burden. As the case for many other complex diseases, early diagnosis is key to prevent irreversible end-organ damages. However, given its mild initial symptoms, T2DM is often diagnosed several years after its onset, leaving half of diabetic individuals undiagnosed. While several classical clinical and genetic biomarkers have been identified, improving early diagnosis by exploring other kinds of omics data remains crucial. In this study, we have combined longitudinal data from two population-based cohorts CoLaus and DESIR (comprising in total 493 incident cases vs 1’360 controls) to identify new or confirm previously implicated metabolomic biomarkers predicting T2DM incidence more than five years ahead of clinical diagnosis. Our longitudinal data have shown robust evidence for valine, leucine, carnitine and glutamic acid being predictive of future conversion to T2DM, and also confirmed to be causal by 2-sample Mendelian randomisation (based on independent data). Interestingly, for valine and leucine a strong reverse causal effect was detected, indicating that the genetic predisposition to T2DM may trigger early changes of these metabolites, which appear well-before any clinical symptoms. These findings indicate that molecular traits linked to the genetic basis of T2DM may be particularly promising early biomarkers.

## 1. Introduction

Type 2 Diabetes Mellitus (T2DM) is a major public health concern and its prevalence is increasing. Almost 500 million individuals are currently affected worldwide by diabetes and almost 700 million may be affected by 2045 [https://www.diabetesatlas.org/en/sections/worldwide-toll-of-diabetes.html]. Diabetes remains among the leading causes of cardiovascular disease, blindness, kidney failure, and lower-limb amputation. By the time T2DM is diagnosed, many individuals have already established end-organ damage including neuropathy, kidney failure and/or premature cardiac or brain atherosclerosis. Diabetes and pre-diabetes are diagnosed by routinely assessed clinical markers (glycaemia and HbA_1_c levels) above a given threshold. Still, agreement between the different markers in diagnosing T2DM is not optimal [1], and their screening capacity for pre-diabetes is low [2]. While these markers are powerful predictors of the disease, they are far from perfect for the identification of individuals who are prone to develop T2DM. Early detection of individuals with high T2DM predisposition is important as non-pharmacological approaches (i.e. lifestyle changes) can reduce substantially (and at a reduced cost) the risk of developing T2DM [3]. Thus, the use of predictive biological tests or clinical scores enabling a more precise identification of individuals at risk is a pressing need so that preventive measures can be applied to limit the increase of T2DM prevalence world-wide. Several predictive scores using clinical data and/or biological markers have been developed (e.g. [4]), but their performance (AUC ∼0.75) is far from optimal. Over 90% positive- and negative predictive value would be clinically relevant, which is achievable for some diseases [5]. Recently, polygenic scores based on cross-sectional T2DM case-control status have been explored and may be of great value to enhance the predictive capacity of the disease onset and to better understand the clinical heterogeneity of T2DM [6]. Since many metabolites and proteins are expected to be altered in pre-diabetic state, using different omics profiles in addition to the classical clinical, biological and genetic risk factors is expected to increase the prediction accuracy.

Several cross-sectional and longitudinal metabolomic studies, focussing on blood samples using targeted approaches, have been initiated to identify candidate biomarkers of pre-diabetes, with a few exceptions employing untargeted approaches (e.g. a cross-sectional study of 115 T2DM individuals [7]). In the population-based cooperative health research of Augsburg (KORA), 140 metabolites were quantified for 4,297 participants and several metabolites altered in pre-diabetic individuals have been identified [8]. Using metabolite-protein network and targeted approaches on serum samples, Wang-Sattler *et al*. identified seven T2DM-related genes associated with these metabolites by multiple interactions with four enzymes. Lysophosphatidylcholine (18:2) and glycine were strong predictors of glucose intolerance, even 7 years before disease onset. These metabolites, in addition to sugar metabolites, acylcarnitines and other aminoacids, have been identified as predictors of T2DM also in the European Prospective Investigation into Cancer and nutrition cohort [9]. More recently, Padberg *et al*. described a metabolic signature that includes glyoxylate associated with T2DM and prediabetic individuals [10]. Wang *et al*, using longitudinal data on 201 incident T2DM cases, identified a signature of five branched-chain and aromatic metabolites for which individuals in the top quartile exhibited a five-fold higher risk to develop T2DM [11]. Particularly, a combination of three amino acids predicted future T2DM, with a more than five-fold higher risk for individuals in top quartile, suggesting that amino acid profiles could aid in diabetes risk assessment. These results were confirmed by a recent meta-analysis from 8 prospective studies on 8’000 individuals, which found a higher risk of T2DM for isoleucine, leucine, valine and phenylalanine [12].

By targeting serum carnitine metabolites on 173 incident T2DM cases among 2’519 patients with coronary artery disease, Strand *et al*. demonstrated that trimethyl-lysine, g-butyrobetaine, as both precursors on free carnitine and palmitoyl-carnitine, predict long-term risk of T2DM independently of traditional risk factors [13].

As another example, shotgun lipidomics was applied in a transversal study on plasma of pre-diabetic mice from different genetic backgrounds and revealed a group of ceramides correlated with glucose tolerance and insulin secretion [14]. These results were interestingly confirmed by quantitative analysis in the plasma of individuals from two population-based prospective cohorts showing that dihydro-ceramides were significantly elevated in the plasma of individuals who will progress to diabetes up to 9 years before disease onset [14]. Other studies have struggled to identify the contribution of individual metabolites and focused more on metabolome-wide prediction [15], which are difficult to replicate.

The previously listed studies provide several important candidate metabolites to benchmark our experimental and modelling setup. Here, we used a subset of the CoLaus study intentionally enriched for T2DM incident cases to maximise discovery power of baseline metabolite levels being associated with developing T2DM at a later follow-up stage. We compared our findings with a similarly sized population-based cohort, DESIR, and also with bidirectional Mendelian randomisation using metabolite- and T2DM QTLs as instruments.

## 2. Methods

### 2.1 The CoLaus study

The CoLaus study (www.CoLaus-psycolaus.ch) is a population-based prospective study based on a single random sample of 6’733 participants from the overall population aged between 35-75 living in Lausanne (10). The baseline survey was conducted between 2003 and 2006. Each participant was extensively phenotyped regarding personal, lifestyle and cardiovascular risk factors; extensive blood and urine characterization was performed, and over 500’000 SNPs were directly genotyped and a further 20.4 million imputed (with r2-hat>0.3). The first follow-up was performed between April 2009 and September 2012; median follow-up time was 5.4 (average 5.6, range 4.5-8.8) years; it included 5’064 participants, and the 5.5-year incidence of T2DM was 6.5%, with 284 incident cases. The second follow-up was performed between May 2014 and April 2017; median follow-up was 10.7 (average 10.9, range 8.8-13.6) years. In this study, we selected 262 T2DM incident cases at the first follow-up and 524 controls matched for sex, age and baseline glucose. For each case, two types of controls were selected: one with a very low risk of T2DM (as assessed by a multivariable T2DM risk score [16]) and one with pre-diabetes (with a high-risk score, but no T2DM at the CoLaus second follow-up). Incident T2DM cases were defined as fasting glucose ≥ 7 mmol/L and/or presence of antidiabetic drug treatment and/or HbA1c ≥ 6.5%. The most important study characteristics are included in Table 1. The study protocols were approved by the Ethical Committee of the Canton de Vaud and all participants provided written informed consent.

**Table 1.** Sample characteristics of the CoLaus and DESIR studies. Yn denotes n years after baseline (e.g. Y3 means a follow-up 3 years after baseline). N stands for sample size. Values are either number (% of total), median [interquartile range] or mean ± standard deviation.

|  | CoLaus |  |  | DESIR |  |  |  |
| --- | --- | --- | --- | --- | --- | --- | --- |
|  | Y0 | Y5 | Y10 | Y0 | Y3 | Y6 | Y9 |
| **N** | 788 | 788 | 673 | 984 | 961 | 932 | 957 |
| **Gender (woman)** | 268 (34.0) | 268 (34.0) | 240 (35.7) | 469 (47.7) | 455 (47.3) | 442 (47.4) | 462 (48.3) |
| **Age (years)** | 56.2 ± 9.7 | 61.8 ± 9.7 | 66.7 ± 9.5 | 48.2 ± 10.1 | 51.2 ± 10.1 | 54.2 ± 10.1 | 57.2 ± 10.1 |
| **Smoking categories (%)** |  |  |  |  |  |  |  |
| **Never** | 283 (35.9) | 280 (36.0) | 237 (38.4) | 484 (49.2) | 474 (49.3) | 456 (48.9) | 473 (49.4) |
| **Former** | 302 (38.3) | 339 (43.6) | 284 (46.0) | 278 (28.3) | 272 (28.3) | 264 (28.3) | 270 (28.2) |
| **Current** | 203 (25.8) | 159 (20.4) | 96 (15.6) | 222 (22.6) | 215 (22.4) | 212 (22.7) | 214 (22.4) |
| **Alcohol consumption (g/week)** | 5 [1-11] | 4 [0-10] | 4 [0-10] | 2.51 [0.14 - 3.29] | 2.54 [0.29 - 4.04] | 2.48 [0.29 - 4.04] | 2.29 [0.29 - 3.52] |
| **Body mass index (kg/m<sup>2</sup>)** | 27.1 ± 4.6 | 27.5 ± 4.9 | 27.8 ± 5.1 | 25.1 ± 4.0 | 25.6 ± 4.4 | 26.0 ± 4.5 | 26.2 ± 4.5 |
| **Waist (cm)** | 94.2 ± 13.1 | 97.1 ± 13.2 | 97.7 ± 13.9 | 84.5 ± 12.1 | 85.9 ± 12.6 | 87.5 ± 12.9 | 88.2 ± 13.1 |
| **HDL (mmol/L)** | 1.53 ± 0.42 | 1.53 ± 0.44 | 1.49 ± 0.46 | 1.61 ± 0.42 | 1.51 ± 0.39 | 1.62 ± 0.40 | 1.47 ± 0.34 |
| **Triglycerides (mmol/L)** | 1.3 [0.9-1.9] | 1.3 [0.9-1.8] | 1.2 [0.9-1.7] | 1.2 [0.7 - 1.5] | 1.3 [0.7 - 1.6] | 1.3 [0.8 - 1.6] | 1.3 [0.8 - 1.5] |
| **Glucose (mmol/L)** | 5.68 ± 0.66 | 6.38 ± 1.13 | 5.98 ± 1.39 | 5.39 ± 0.57 | 5.53 ± 0.84 | 5.63 ± 1.04 | 5.67 ± 1.06 |
| **Insulin (μU/mL)** | 8.2 [5.1-12.4] | 8.3 [5.2-13.5] | 9.8 [5.9-14.8] | 7.1 [4.2 - 8.3] | 7.0 [4.2 - 8.4] | 7.5 [3.9 - 9.0] | 7.3 [4.1 - 8.7] |
| **HOMA2-IR** | 2.1 [1.3-3.3] | 2.4 [1.4-4.0] | 2.6 [1.4-4.2] | 1.1 [0.7 - 1.3] | 1.1 [0.7 - 1.3] | 1.2 [0.7 - 1.5] | 1.2 [0.7 - 1.4] |

### 2.2 The DESIR cohort

The prospective D.E.S.I.R. cohort is a nine-year follow up study of 2,391 middle-aged European ancestry participants [17-19]. We analysed participants from a case-cohort design embedded within the larger cohort that includes 231 cases of incident T2DM and 836 participants randomly sampled from the entire cohort. Baseline and follow-up clinical characteristics of participants included in the training population are shown in [4] (see Table 1). T2DM was defined using one of the following criteria: use of glucose lowering medication, fasting plasma glucose [FG] ≥7 mmol/L, or glycated hemoglobin A1c [HbA_1c_] ≥6.5% (48 mmol/mol). Clinical and biological evaluations were performed at inclusion and after three, six, and nine years, as previously described [20]. All participants provided written informed consent and the study protocol was approved by the Ethics Committee for the Protection of Subjects for Biomedical Research of Bicêtre Hospital, France. Metabolites measurements have been described elsewhere in full details [20].

### 2.3 Targeted Metabolomics analysis

Plasma and urine samples collected at the baseline of the CoLaus cohort were processed for targeted metabolomics analysis as described elsewhere [21]. Briefly, metabolites were extracted from 100 μL of plasma or urine samples and Quality Control (QC) samples using a cold methanol-ethanol solvent mixture in a 1:1 ratio. After centrifugation at 14’000 rpm for 15 minutes, supernatant was recovered, evaporated and resuspended in 100μL (for plasma) or 200μL (for urine) of H2O:MeOH (9:1). 5μL of the samples were analyzed by LC-MRM/MS on a hybrid triple quadrupole-linear ion trap QqQ_LIT_ (Qtrap 5500, Sciex) hyphenated to a LC Dionex Ultimate 3000 (Dionex, Thermo Scientific). Analyses were performed in positive and negative electrospray ionization using a TurboV ion source. The chromatographic separation was performed on a Kinetex column C18 (100×2.1 mm, 2.6 μm). The mobile phases were constituted by A: H_2_O with 0.1% FA and B: ACN with 0.1% FA for the positive mode. In the negative mode, the mobile phases were constituted by A: ammonium fluoride 0.5 mM in H _2_O and B: ammonium fluoride 0.5 mM in ACN. The linear gradient program was 0-1.5 min 2%B, 1.5-15 min up to 98%B, 15-17 min held at 98% B, 17.5 min down to 2%B at a flow rate of 250 μL/min.

The MRM/MS method included 299 and 284 transitions in positive and negative mode respectively, corresponding to 583 endogenous metabolites. For each biological matrix, the 786 samples were prepared and analyzed in 8 batches. In order to monitor the signal drift and system performance over time, and to avoid repeated thawing-freezing cycles of the study samples, quality control (QCs) surrogate samples were used. These QC samples were prepared in the same way and at the same time of the study samples from aliquots of a pool of human plasma or urine that was the same for all the analytical batches. QC samples were injected every 8 samples in both positive and negative modes.

The MS instrument was controlled by Analyst software v.1.6.2 (AB Sciex). Peak integration was performed with MultiQuant software v.3.0 (AB Sciex). The integration algorithm was MQ4 with a Gaussian smoothing of a half-width equal to 1.5 points. For plasma samples, the analysis was narrowed to the 124 and 48 metabolites that were detected in all samples with a noise percentage of 80% and a gaussian peak shape, in positive and negative modes, respectively. For urine samples, we detected 124 and 77 metabolites in positive and negative modes, respectively. To correct for batch effect, raw data were normalized with the dbnorm package [22], by using the *ber* model.

### 2.4 Statistical approaches

We performed logistic- and linear regression analysis to test for association between baseline metabolite levels and T2DM incidence and glucose level changes, respectively. We included the following covariates: family history of diabetes, smoking status, body-mass index (BMI), HDL cholesterol, triglycerides, insulin, glucose and HOMA measure at baseline. Since none of our association P-values passed strict Bonferroni correction for multiple testing (P<0.05/172, we declared P-values below 0.01 as suggestively significant.

### 2.5 Bi-direction Mendelian Randomization

For each metabolite associated with glucose, we performed two-sample bidirectional Mendelian randomization (MR), an instrumental variable method to distinguish correlation from causation in observational data [23]. The idea of MR is to use genetic variants as instrumental variables to attempt causal inference about the effect of modifiable risk factors, which can overcome some types of confounding and reverse causation.

We tested whether genetically predicted levels of a particular metabolite affect the risk for elevated glucose and type 2 diabetes and whether genetically increased risk of type 2 diabetes or elevated glucose is associated with circulating levels of a particular metabolite. The associations between the instrumental variables and the exposure and the outcome are estimated from independent studies, either the metabolite GWAS or fasting glucose/T2DM GWAS.

For each metabolite associated with glucose, we performed two-sample bidirectional MR. We tested whether genetically varying levels of a particular metabolite affect the risk for elevated glucose and T2DM (we call this MR) and whether genetically increased risk of T2DM or elevated glucose is associated with circulating levels of a particular metabolite (we call this reverse MR). The associations between the instrumental variables and the exposure and the outcome are estimated from independent studies. We used summary statistics from the UKBB for glucose and those published by the DIAGRAM Consortium for T2DM [24]. The metabolite data are from a large GWAS performed on 7,824 adult individuals from European population studies.

## 3. Results

### 3.1 Study characteristics

Selected basic features of the CoLaus study are listed in Table 1. We selected 788 participants, including 263 T2DM incident cases at the first follow-up and 525 controls matched for sex, age and baseline glucose. Summary data are expressed either as counts (and percentage) for categorical variables and as median [interquartile range] or mean ± standard deviation for continuous variables.

### 3.2 Metabolites association scan

Based on quality criteria such as sensitivity and peak shape, we detected 172 urine and plasma metabolites (MS) in the 788 selected CoLaus participants. The analytical samples were collected at the baseline and included 525 participants without diabetes and 263 participants who became diabetic over the following 10 years. For each metabolite, we ran association scans to find those correlated to change in diabetes or glucose levels. All the results are based on the first follow-up which is the most predictive analysis. Indeed, when we compared the effects estimated in the first and second follow-up we observed significant weaker effects in the second follow-up (*P*_t-test_ =1.38 ×10^−05^, see Figure1).

**Figure1.**
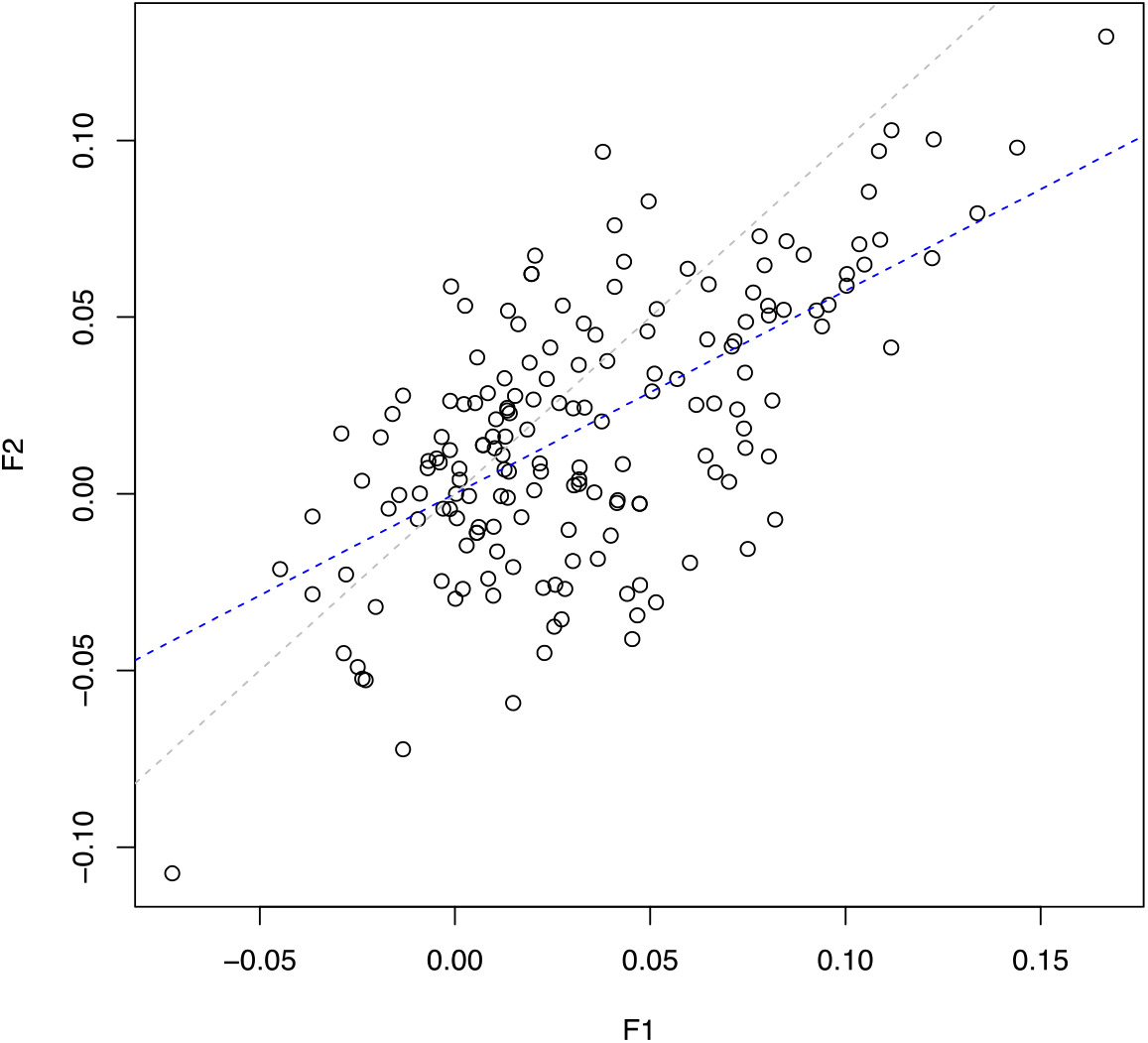
Linear relationship between the effects estimated in the first (F1) and second (F2) follow-up. The blue and grey lines represent the regression and the identity line respectively.

The metabolome-wide association scan revealed seven metabolites associated with glucose change at suggestively significant level (P<0.01) in the CoLaus study, see Table 2. When we meta-analysed metabolome-wide results from the CoLaus and DESIR studies, we similarly found leucine and four additional suggestively significant (P<0.01) metabolites associated with glucose change (Table 3).

**Table 2.** Metabolites associated with glucose change in the CoLaus cohort. For each metabolite we report its effect size on glucose in the CoLaus and DESIR cohorts, the combined P-value and the forward and reverse causal effect on glucose and T2DM estimated by Mendelian Randomisation. “X” indicates missing value, i.e. when the metabolite was not available for the respective analysis. The ID for the Human Metabolome Database is indicated for each metabolite.

| Metabolite | CoLaus |  | DESIR |  | CoLaus+DESIR | Metabolite -> Glucose MR |  | Glucose -> metabolite MR |  | Metabolite -> T2DM MR |  | T2DM -> Metabolite MR |  |
| --- | --- | --- | --- | --- | --- | --- | --- | --- | --- | --- | --- | --- | --- |
|  | effect | P-value | effect | P-value | P-value | effect | P-value | effect | P-value | effect | P-value | effect | P-value |
| **Valine**<br>(HMDB0000883) | 0.167 | 9.00E-04 | 0.005 | 0.853 | 0.069 | 0.000 | 1.000 | 0.104 | 0.023 | 0.023 | 0.008 | 0.115 | 0.003 |
| **Leucine**<br>(HMDB0000687) | 0.134 | 0.0021 | 0.046 | 0.158 | 2.90E-03 | 0.002 | 0.372 | 0.042 | 0.360 | 0.007 | 0.011 | 0.159 | 5.79E-05 |
| **Sarcosine**<br>(HMDB0000271) | 0.144 | 0.0039 | 0.014 | 0.581 | 0.076 | x | x | x | x | x | x | x | x |
| **4-Hydroxyproline**<br>(HMDB00006055) | 0.122 | 0.0042 | -0.039 | 0.116 | 0.94 | x | x | x | x | x | x | x | x |
| **Glutamic acid**<br>(HMDB00148) | 0.105 | 0.009 | -0.038 | 0.185 | 0.656 | 0.01 | 0.147 | 0.069 | 0.134 | 0.012 | 0.12 | 0.18 | 2.84E-06 |
| **Homoserine**<br>(HMDB0000719) | 0.123 | 0.0094 | x | x | x | x | x | x | x | x | x | x | x |

**Table 3.**
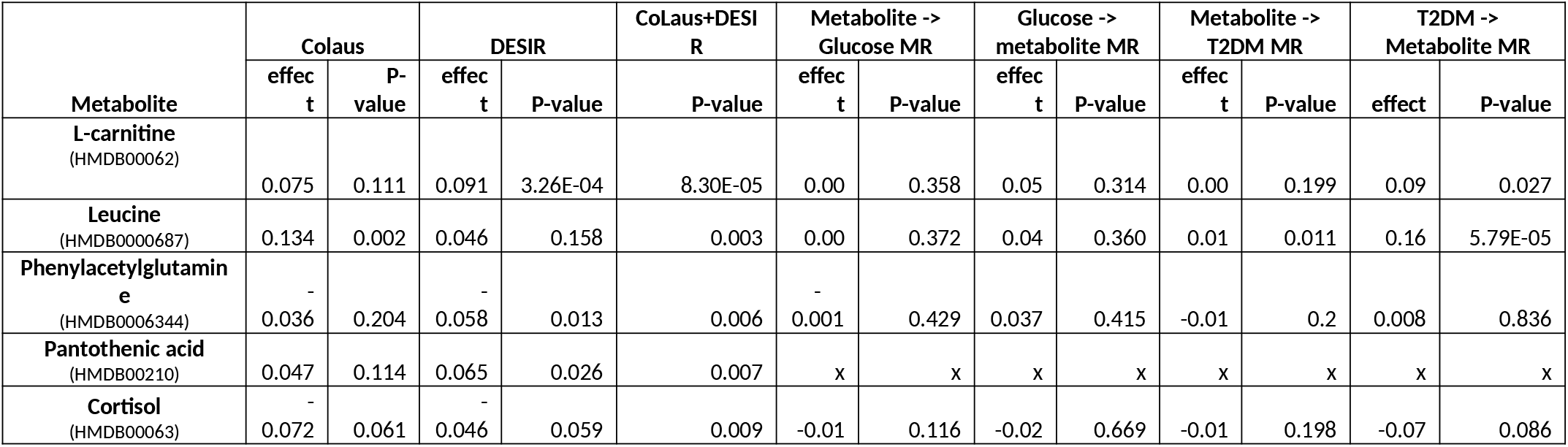
Additional metabolites found significantly associated with glucose change after combining CoLaus and DESIR Cohort results. For each metabolite we report its effect size on glucose in CoLaus and DESIR cohorts, the combined P-value and the forward and reverse causal effect on glucose and T2DM estimated by Mendelian Randomisation. The ID for the Human Metabolome Database is indicated for each metabolite.

Finally, three out of the seven metabolites associated with glucose were testable with MR. Interestingly, T2DM showed a significant causal effect on valine (P = 0.003), leucine (P = 5.8 × 10^−05^) and glutamate (P= 2.8 × 10^−06^).

## 4. Discussion

Using mass-spectrometry targeted metabolomics analysis, we identified a panel of metabolites whose levels are associated with glucose changes before the onset of T2DM in the CoLaus Cohort. We replicated our findings in an independent study (from DESIR), which reassuringly revealed five metabolites with combined P-value below 0.01. While we focused our analysis on predicting T2DM for a 5-years follow-up period, we observed that the effect of the potentially predictive metabolites diminished over time. This is unsurprising as risk factors change over time, hence more and more unknowns contributing to diabetes conversion accumulate with time. We have noticed, furthermore, that more than 80% (140/172) of the metabolite effects are positive, meaning that generally an increased level of metabolites represents a risk factor for diabetes. This observation needs to be considered with caution, since it might be due to a latent diabetes-associated confounding factor, which is linked to overall metabolite concentration. This explanation is rather unlikely since we accounted for metabolomic principal components in all association scan.

The confirmed metabolites have been repeatedly supported by various types of evidence in both human and model organisms. Individuals with obesity and T2DM have elevated levels of branched-chain amino acids (leucine, isoleucine and valine) [11, 25-28]. Such changes are already present before the onset of diabetes [11, 12] and their causal role is believed to be exerted via the modulation of the mTOR pathway. Increased leucine levels can lead to insulin resistance via activation of the TORC1 pathway, with induction of beta cell proliferation and insulin secretion [11, 29] and disruption of insulin signal in skeletal muscle [30]. On the other hand, insulin resistance enhances protein catabolism in skeletal muscle, which can increase the release of branched-chain amino acids [31]. Moreover, hyperglycemia negatively correlates with adipose tissue expression of genes involved in branched-chain amino acid oxidation, which can further contribute to raise the levels of BCAA [31]. Thus, so far it remains unclear whether the observed BCAA changes are only a consequence of hyperglycemia or if they have a causative role in the development of T2DM.

Another metabolite which is displaying a significant association with glucose changes in the CoLaus cohort is glutamic acid, although this is not replicated in the DESIR study. A link between increased glutamate levels and insulin resistance traits was already observed in multiple cohorts [26, 32], and more recently, a meta-analysis conducted in 18 prospective studies highlighted glutamate as positively correlated with T2DM [33]. Glutamate is a glucogenic aminoacid, which can enter into the Krebs cycle through its conversion to α-ketoglutarate. In addition, it can favour gluconeogenesis by increasing the transamination of pyruvate to alanine [34], and can directly stimulate glucagon release from pancreatic α-cells [35]. However, its possible causal role remains controversial and several reports suggest that it is rather a reduced ratio between glutamine, a glutamate derivative, and glutamate itself, that is informative of metabolic risk [32, 36].

In our study, free carnitine, cortisol, phenylacetylglutamine and pantothenic acid also appeared as significantly associated to the development of T2DM, although only when our data were combined with the French cohort (CoLaus+DESIR). Carnitine esterification with fatty acids is required for the shuttling of the latter into the mitochondria for fatty acid oxidation. Lower levels of free carnitine were reported in diabetic individuals [13, 37, 38], while changes of acylcarnitines and/or carnitine precursors were highlighted in several studies as indicators of prediabetes/T2DM [8, 9, 27], although data are not always consistent. Our targeted LC-MS method included several of these acylcarnitines, such as acetylcarnitine, propionylcarnitine, and isovalerylcarnitine, but no significant association was found with subsequent development of T2DM.

Cortisol dysregulation has been linked to T2DM in cross-sectional and longitudinal studies [39, 40]. More specifically, diabetic individuals present a flattened diurnal cortisol curve compared to non-diabetic ones [41], with lower morning and higher afternoon and evening concentrations [42]. Interestingly, high levels of evening cortisol were also shown to be predictive of T2DM development in an occupational cohort [43]. Even though the mechanisms underlying this association are not completely understood, cortisol contributes to many metabolic processes that can potentially perturb glucose homeostasis [44]. Cortisol has a major role in raising glucose levels through gluconeogenesis activation. Moreover, it induces lipolysis, thus increasing the release of free fatty acids that may favour the impairment of glucose uptake. Of note, diabetes is considered a common complication in clinical states characterized by prolonged hypercortisolaemia, such as in Cushing disease [45]. However, the causality of this association remains to be determined. Phenylacetylglutamine is a nitrogenous metabolite almost exclusively derived from the gut microbiota conversion of phenylalanine [46]. Its accumulation is known to occur in uraemia and was shown to be increased in type 2 diabetic patients, particularly in association with renal damage [47, 48]. Other findings with variable degree of evidence in our study have also solid corroborating literature. Sarcosine (N-methylglycine) is an intermediate and by-product in glycine synthesis and has been found to be a moderately strong (OR=1.3) predictor of T2DM incidence [49]. More specifically, the addition of urine sarcosine to other established predictors of incident T2DM was shown to improve model performance and T2DM risk prediction in a cohort of 4’164 patients with suspected stable angina pectoris. Diabetic individuals have higher circulating proline levels and, moreover, proline-induced insulin transcription impairment may contribute to the β-cell dysfunction observed in T2DM [50].

### 4.1 Causal inference

Drawing causal inference is extremely difficult. While most human studies are observational and cross-sectional predominantly, only correlations are calculated between a disease status and the levels of various predictors. Such a simple measure cannot tease apart forward-, reverse causation or confounding. Longitudinal studies provide more specific directional link (called Granger causality) between a potential biomarker and disease outcomes. With its roots in differential equation modelling, an association between the baseline level of a predictor and the change of the outcome over time may imply a causal relationship. An orthogonal axis of evidence for causality can be provided by Mendelian randomization (MR), where exposure-associated genetic markers act as instruments to tease out the causal relationship between a potential risk factor and an outcome [23]. Interventional studies, due to their intrusive nature are performed mostly in model organisms and can help triangulating causal evidence. Comparisons between the different approaches for causality have been very scarce due to the little overlap between the respective scientific communities. A pioneering work [51] in this aspect has shown good agreement between disease-to-biomarker MR results, observational correlation and longitudinal associations. However, their longitudinal association included exposure (adiposity) change regressed on outcome (metabolite level) change, which implies no directionality and is not the intuitive way to perform such analysis. Among the metabolites that we identified here as early biomarkers of T2D, we have shown that leucine has bidirectional causal relationship with diabetes, with a reverse (diabetes to metabolite) causal effect that seems to be stronger. Interestingly, (predisposition to) diabetes has a significant causal effect also on glutamate and valine, while their direct effect on diabetes is not significant. Hence small molecules targeting these metabolites may be more effective for treating downstream organ damage of T2DM, such as cardiovascular disease, neuropathies or nephropathy. In line with this hypothesis, glutamate accumulation in the retina can cause neurotoxicity and the development of diabetic retinopathy [52, 53], even though the actual connection between plasma and retinal glutamate levels remains to be assessed [36].

### 4.2 Strengths and limitations

Our study has numerous strengths. First, is our use of two well characterized prospective cohorts (one for replication), whose participants have been followed longitudinally for more than ten years. This approach allowed us to investigate potential biomarkers in blood samples collected when individuals were still free of diabetes. Second, the robustness of our targeted methods and of our results is evidenced by the fact that we confirm many previous findings. Third, we triangulate evidence by combining these longitudinal association results with other causal inference techniques, such as Mendelian Randomisation.

The major limitation of this study is the relatively low number of incident diabetes cases that we could analyse, which prohibited us from new discoveries with unequivocal statistical evidence. In the light of these findings, we recommend future research focussing more on untargeted metabolomic approaches better exploring the vast space of metabolite species and the investigation of other omics biomarkers in parallel.

### 4.2 Conclusions

Our study has confirmed most of the identified-to-date metabolites in a medium-sized longitudinal population-based study (enriched for incident cases) and provided complementary evidence from bi-directional Mendelian randomisation. However, the quest for early metabolic biomarkers predicting the development of T2DM require more research effort including larger studies in order to understand the potentially minute contributions of many circulating metabolites.

## Data Availability

The individual data from CoLAus and DESIR cohorts are not publicly available.

## Acknowledgements

The study was supported by the Swiss National Science Foundation (#32473B-166450; FN 32003B_173092, and #31003A_182420).

## Notes

### Competing Interest Statement

The authors have declared no competing interest.

### Author Declarations

CoLaus cohort: The study protocols were approved by the Ethical Committee of the Canton de Vaud and all participants provided written informed consent. DESIR cohort: All participants provided written informed consent and the study protocol was approved by the Ethics Committee for the Protection of Subjects for Biomedical Research of Bicentre Hospital, France.

## References

1. Tucker, L.A., Limited Agreement between Classifications of Diabetes and Prediabetes Resulting from the OGTT, Hemoglobin A1c, and Fasting Glucose Tests in 7412 US Adults. Journal of Clinical Medicine, 2020. 9(7).

2. Serdar, M.A., et al., An Assessment of HbA1c in Diabetes Mellitus and Pre-diabetes Diagnosis: a Multi-centered Data Mining Study. Appl Biochem Biotechnol, 2020. 190(1): p. 44–56.

3. Kyi, M., et al., Early Intervention for Diabetes in Medical and Surgical Inpatients Decreases Hyperglycemia and Hospital-Acquired Infections: A Cluster Randomized Trial. Diabetes Care, 2019. 42(5): p. 832–840.

4. Yengo, L., et al., Impact of statistical models on the prediction of type 2 diabetes using non-targeted metabolomics profiling. Molecular Metabolism, 2016. 5(10): p. 918–925.

5. Fenske, W., et al., A Copeptin-Based Approach in the Diagnosis of Diabetes Insipidus. N Engl J Med, 2018. 379(5): p. 428–439.

6. Udler, M.S., et al., Genetic Risk Scores for Diabetes Diagnosis and Precision Medicine. Endocr Rev, 2019. 40(6): p. 1500–1520.

7. Menni, C., et al., Biomarkers for Type 2 Diabetes and Impaired Fasting Glucose Using a Nontargeted Metabolomics Approach. Diabetes, 2013. 62(12): p. 4270–4276.

8. Wang-Sattler, R., et al., Novel biomarkers for pre-diabetes identified by metabolomics. Molecular Systems Biology, 2012. 8.

9. Floegel, A., et al., Identification of Serum Metabolites Associated With Risk of Type 2 Diabetes Using a Targeted Metabolomic Approach. Diabetes, 2013. 62(2): p. 639– 648.

10. Padberg, I., et al., A New Metabolomic Signature in Type-2 Diabetes Mellitus and Its Pathophysiology. Plos One, 2014. 9(1).

11. Wang, T.J., et al., Metabolite profiles and the risk of developing diabetes. Nat Med, 2011. 17(4): p. 448–53.

12. Guasch-Ferre, M., et al., Metabolomics in Prediabetes and Diabetes: A Systematic Review and Meta-analysis. Diabetes Care, 2016. 39(5): p. 833–46.

13. Strand, E., et al., Serum Carnitine Metabolites and Incident Type 2 Diabetes Mellitus in Patients With Suspected Stable Angina Pectoris. J Clin Endocrinol Metab, 2018. 103(3): p. 1033–1041.

14. Wigger, L., et al., Plasma Dihydroceramides Are Diabetes Susceptibility Biomarker Candidates in Mice and Humans. Cell Rep, 2017. 18(9): p. 2269–2279.

15. Liu, J., et al., Metabolomics based markers predict type 2 diabetes in a 14-year follow-up study. Metabolomics, 2017. 13(9): p. 104.

16. Schmid, R., et al., Validation of 7 type 2 diabetes mellitus risk scores in a population-based cohort: CoLaus study. Arch Intern Med, 2012. 172(2): p. 188–9.

17. Balkau, B., An epidemiologic survey from a network of French health examination centres. (DESIR: Epidemiologic data on the insulin resistance syndrome). Revue D Epidemiologie Et De Sante Publique, 1996. 44(4): p. 373–375.

18. Bonnet, F., et al., Parental history of type 2 diabetes, TCF7L2 variant and lower insulin secretion are associated with incident hypertension. Data from the DESIR and RISC cohorts. Diabetologia, 2013. 56(11): p. 2414–2423.

19. Vaxillaire, M., et al., Type 2 diabetes-related genetic risk scores associated with variations in fasting plasma glucose and development of impaired glucose homeostasis in the prospective DESIR study. Diabetologia, 2014. 57(8): p. 1601– 1610.

20. Balkau, B., et al., Proposed criteria for the diagnosis of diabetes: Evidence from a French epidemiological study (DESIR). Diabetes & Metabolism, 1997. 23(5): p. 428– 434.

21. Domingo-Almenara, X., et al., XCMS-MRM and METLIN-MRM: a cloud library and public resource for targeted analysis of small molecules. Nat Methods, 2018. 15(9): p. 681–684.

22. Bararpour, N., et al., Visualization and normalization of drift effect across batches in metabolome-wide association studies. bioRxiv, 2020.

23. Smith, G.D. and S. Ebrahim, ‘Mendelian randomization’: can genetic epidemiology contribute to understanding environmental determinants of disease? International Journal of Epidemiology, 2003. 32(1): p. 1–22.

24. Scott, R.A., et al., An Expanded Genome-Wide Association Study of Type 2 Diabetes in Europeans. Diabetes, 2017. 66(11): p. 2888–2902.

25. Chen, X. and W. Yang, Branched-chain amino acids and the association with type 2 diabetes. J Diabetes Investig, 2015. 6(4): p. 369–70.

26. Ferrannini, E., et al., Early metabolic markers of the development of dysglycemia and type 2 diabetes and their physiological significance. Diabetes, 2013. 62(5): p. 1730–7.

27. Palmer, N.D., et al., Metabolomic profile associated with insulin resistance and conversion to diabetes in the Insulin Resistance Atherosclerosis Study. J Clin Endocrinol Metab, 2015. 100(3): p. E463–8.

28. Wurtz, P., et al., Circulating metabolite predictors of glycemia in middle-aged men and women. Diabetes Care, 2012. 35(8): p. 1749–56.

29. Melnik, B.C., Leucine signaling in the pathogenesis of type 2 diabetes and obesity. World J Diabetes, 2012. 3(3): p. 38–53.

30. Krebs, M., et al., Mechanism of amino acid-induced skeletal muscle insulin resistance in humans. Diabetes, 2002. 51(3): p. 599–605.

31. Stancakova, A., et al., Hyperglycemia and a common variant of GCKR are associated with the levels of eight amino acids in 9,369 Finnish men. Diabetes, 2012. 61(7): p. 1895–902.

32. Cheng, S., et al., Metabolite profiling identifies pathways associated with metabolic risk in humans. Circulation, 2012. 125(18): p. 2222–31.

33. Sun, Y., et al., Metabolomics Signatures in Type 2 Diabetes: A Systematic Review and Integrative Analysis. J Clin Endocrinol Metab, 2020. 105(4).

34. Newgard, C.B., et al., A Branched-Chain Amino Acid-Related Metabolic Signature that Differentiates Obese and Lean Humans and Contributes to Insulin Resistance (vol 9, pg 311, 2009). Cell Metabolism, 2009. 9(6): p. 565–566.

35. Cabrera, O., et al., Glutamate is a positive autocrine signal for glucagon release. Cell Metabolism, 2008. 7(6): p. 545–554.

36. Rhee, S.Y., et al., Plasma glutamine and glutamic acid are potential biomarkers for predicting diabetic retinopathy. Metabolomics, 2018. 14(7): p. 89.

37. Poorabbas, A., et al., Determination of free L-carnitine levels in type II diabetic women with and without complications. Eur J Clin Nutr, 2007. 61(7): p. 892–5.

38. Ringseis, R., J. Keller, and K. Eder, Role of carnitine in the regulation of glucose homeostasis and insulin sensitivity: evidence from in vivo and in vitro studies with carnitine supplementation and carnitine deficiency. Eur J Nutr, 2012. 51(1): p. 1–18.

39. Joseph, J.J. and S.H. Golden, Cortisol dysregulation: the bidirectional link between stress, depression, and type 2 diabetes mellitus. Annals of the New York Academy of Sciences, 2017. 1391(1): p. 20–34.

40. Papandreou, C., et al., Plasma metabolites predict both insulin resistance and incident type 2 diabetes: a metabolomics approach within the Prevencion con Dieta Mediterranea (PREDIMED) study. Am J Clin Nutr, 2019. 109(3): p. 626–634.

41. Bruehl, H., O.T. Wolf, and A. Convit, A blunted cortisol awakening response and hippocampal atrophy in type 2 diabetes mellitus. Psychoneuroendocrinology, 2009. 34(6): p. 815–21.

42. Lederbogen, F., et al., Flattened circadian cortisol rhythm in type 2 diabetes. Exp Clin Endocrinol Diabetes, 2011. 119(9): p. 573–5.

43. Hackett, R.A., et al., Diurnal Cortisol Patterns, Future Diabetes, and Impaired Glucose Metabolism in the Whitehall II Cohort Study. J Clin Endocrinol Metab, 2016. 101(2): p. 619–25.

44. Di Dalmazi, G., et al., Glucocorticoids and type 2 diabetes: from physiology to pathology. J Nutr Metab, 2012. 2012: p. 525093.

45. Clayton, R.N., et al., Mortality and morbidity in Cushing’s disease over 50 years in Stoke-on-Trent, UK: audit and meta-analysis of literature. J Clin Endocrinol Metab, 2011. 96(3): p. 632–42.

46. Mair, P., et al., [Infusion or repetitive bolus injection? A clinical study of midazolam/fentanyl and diazepam/fentanyl combination anesthesia in neurosurgical operations]. Anasth Intensivther Notfallmed, 1990. 25 Suppl 1: p. 34–8.

47. Barrios, C., et al., Gut-Microbiota-Metabolite Axis in Early Renal Function Decline. PLoS One, 2015. 10(8): p. e0134311.

48. Suhre, K., et al., Metabolic footprint of diabetes: a multiplatform metabolomics study in an epidemiological setting. PLoS One, 2010. 5(11): p. e13953.

49. Svingen, G.F.T., et al., Prospective Associations of Systemic and Urinary Choline Metabolites with Incident Type 2 Diabetes. Clinical Chemistry, 2016. 62(5): p. 755– 765.

50. Liu, Z., et al., Chronic Exposure to Proline Causes Aminoacidotoxicity and Impaired Beta-Cell Function: Studies In Vitro. Rev Diabet Stud, 2016. 13(1): p. 66–78.

51. Wurtz, P., et al., Metabolic Signatures of Adiposity in Young Adults: Mendelian Randomization Analysis and Effects of Weight Change. Plos Medicine, 2014. 11(12).

52. Kowluru, R.A., et al., Retinal glutamate in diabetes and effect of antioxidants. Neurochem Int, 2001. 38(5): p. 385–90.

53. Li, Q. and D.G. Puro, Diabetes-induced dysfunction of the glutamate transporter in retinal Muller cells. Invest Ophthalmol Vis Sci, 2002. 43(9): p. 3109–16.

